# Induction of type I and III interferons by viral and endogenous stimuli in systemic sclerosis

**DOI:** 10.1101/2025.09.23.25336432

**Authors:** Shamisa Adeli, Össur Ingi Emilsson, Erik Hellbacher, Karin Hjorton, Paul Runeson, Anastasios Christias, Per M Hellström, Johan Rönnelid, Lars Rönnblom, Dag Leonard, Tomas Hansen, Andrei Malinovschi, Maija-Leena Eloranta

**Affiliations:** Department of Medical Sciences, Rheumatology, Uppsala University, Uppsala, Sweden; Department of Medical Sciences, Respiratory, allergy and sleep research, Uppsala University, Uppsala, Sweden; Faculty of Medicine, University of Iceland, Reykjavik, Iceland; Department of Medical Sciences, Gastroenterology/Hepatology, Uppsala University, Uppsala, Sweden; Department of Immunology, Genetics and Pathology, Uppsala University, Uppsala, Sweden; Department of Surgical Sciences, Radiology and Radiological Image Analysis, Uppsala University, Uppsala, Sweden; Department of Medical Sciences, Clinical Physiology, Uppsala University, Uppsala, Sweden

**Keywords:** systemic sclerosis, type I interferon, type III interferon, Toll-like receptors, immune complexes, herpes simplex virus, plasmacytoid dendritic cells

## Abstract

**Objective:** The interferon (IFN) system is activated in systemic sclerosis (SSc), but the driving mechanisms remain unclear. We asked whether type I and III IFN responses to Toll-like receptor (TLR)-7/8/9 stimulation of leukocytes from patients with SSc differ from healthy individuals, and if the IFN production is associated with clinical features.

**Methods:** Peripheral blood mononuclear cells (PBMCs), monocyte-depleted PBMCs, and monocytes were prepared from 45 SSc patients and 47 healthy controls. Cells were stimulated with RNA-containing immune complexes (RNA-IC), an RNA-oligonucleotide (ORN8L), or inactivated herpes simplex virus (HSV) targeting TLR7, TLR8, and TLR9, respectively. IFN-α, -β, -λ1 and -λ2 levels were measured by immunoassays. IFN-α producing cells were analyzed by flow cytometry.

**Results:** SSc-PBMCs produced type I and III IFNs in response to all three stimuli, with HSV inducing the strongest response. Compared to controls, SSc-PBMCs produced less IFN-α (p<0.02), while IFN-β levels were higher in HSV-stimulated SSc-monocytes (342 vs. 59.9 pg/ml, p=0.041). Expression of IFN-λ1/2 was lower than type I IFNs. The IFN responses to TLR7/8 stimulation increased in PBMCs in the presence of IFN-α (priming). Strong HSV-induced IFN-α production was associated with diffuse cutaneous SSc, anti-RNA-polymerase III autoantibodies, and interstitial lung disease (ILD).

**Conclusions:** Leukocytes from SSc patients generally have a reduced IFN-producing capacity, except for virus-induced IFN-β production by monocytes. However, type I IFN priming enhanced the IFN response to TLR-7/8 stimulation, suggesting that viral infections may amplify IFN synthesis in response to endogenous TLR activators, that might aggravate the SSc disease process including development of ILD.

## Introduction

Systemic sclerosis (SSc) is an autoimmune rheumatic disease characterized by vasculopathy, inflammation and fibrosis (1, 2). SSc is subclassified into two major subsets: diffuse cutaneous (dcSSc) and limited cutaneous (lcSSc) (1) but the clinical presentation is heterogenous, with cardiopulmonary manifestations, including interstitial lung disease (ILD), being the most severe. A wide range of antinuclear autoantibodies has been identified in SSc, e.g., anti-topoisomerase (topo I) and anti-RNA polymerase III (RNApol III), typically associated with dcSSc, whereas anti-centromere antibodies are generally linked to lcSSc (3).

The etiopathogenesis of SSc is not fully elucidated, but activation of the type I interferon (IFN) system has been implicated as a central contributor to the disease process (4–6). Type I IFNs (especially IFN-α/β) are crucial for antiviral defense, but can be deleterious when dysregulated, due to their strong immunomodulatory effects (7, 8). An early study established a connection between an activated type I IFN system and SSc, demonstrating that a majority of SSc patients expressed IFN-inducible genes (i.e. an IFN-signature), later confirmed by multiple studies (4, 9, 10). Furthermore, administration of IFN-α in a clinical trial was associated with declined lung function in SSc patients (11). Previously, we demonstrated that serum from SSc patients containing anti-RNP or anti-SSA autoantibodies, combined with RNA-containing material from dying cells, stimulated IFN-α production in peripheral blood mononuclear cells (PBMCs) from healthy donors (12).

Such an aberrant IFN-α response is presumably driven by innate immune sensors, including Toll-like receptors (TLRs) (13). Endosomal TLR7 and TLR9 recognize single-stranded RNA and double-stranded DNA, respectively, originating either from microbes or endogenous sources. The latter could lead to immune complex (ICs) formation and chronic activation of type I IFN system, as described in systemic lupus erythematosus (SLE) (14). We have previously shown that plasmacytoid dendritic cells (pDCs), which express both TLR7 and -9, are potent producers of IFN-α in healthy individuals (15). Moreover, TLR8 expression was detected in pDCs from SSc patients, and the stimulation with a TLR8 agonist induced IFN-α production, unlike in pDCs from healthy donors (16).

Type III IFNs (IFN-λ1-3, also known as IL28A-B and IL29) play an important role in the antimicrobial defence at epithelial surfaces. They are produced by epithelial and myeloid cells, as well as by pDCs following activation via TLR7 and -9 (17, 18). IFN-λs signal through a distinct receptor with a restricted spatial expression, resulting in more localized action (19).

To elucidate the contribution of different types of IFNs to the IFN signature in SSc, we investigated the production of IFN-α, IFN-β (type I) and IFN-λ1/2 (type III) in response to endosomal TLR stimulation in blood leukocytes from patients with SSc and healthy controls (HCs), and assessed the association between IFN levels, autoantibodies, and clinical parameters.

## Methods

### Patients and healthy controls

Forty-five patients (36 females and 9 males), fulfilling the 2013 ACR/EULAR classification criteria for SSc (20) were consecutively enrolled at the Rheumatology outpatient clinic, Uppsala University Hospital. Demographic and clinical data are shown in table 1 and treatment in supplementary table 1. Definitions of the clinical manifestations are presented in the supplementary method file. Forty-seven healthy blood donors (34 females and 13 males; median age 53 years, range 22-69) were recruited at the Dept. of Clinical immunology and Transfusion medicine, Uppsala University Hospital. The study was approved by the national Ethics Committee and written informed consent was obtained from all participants.

**Table 1.**
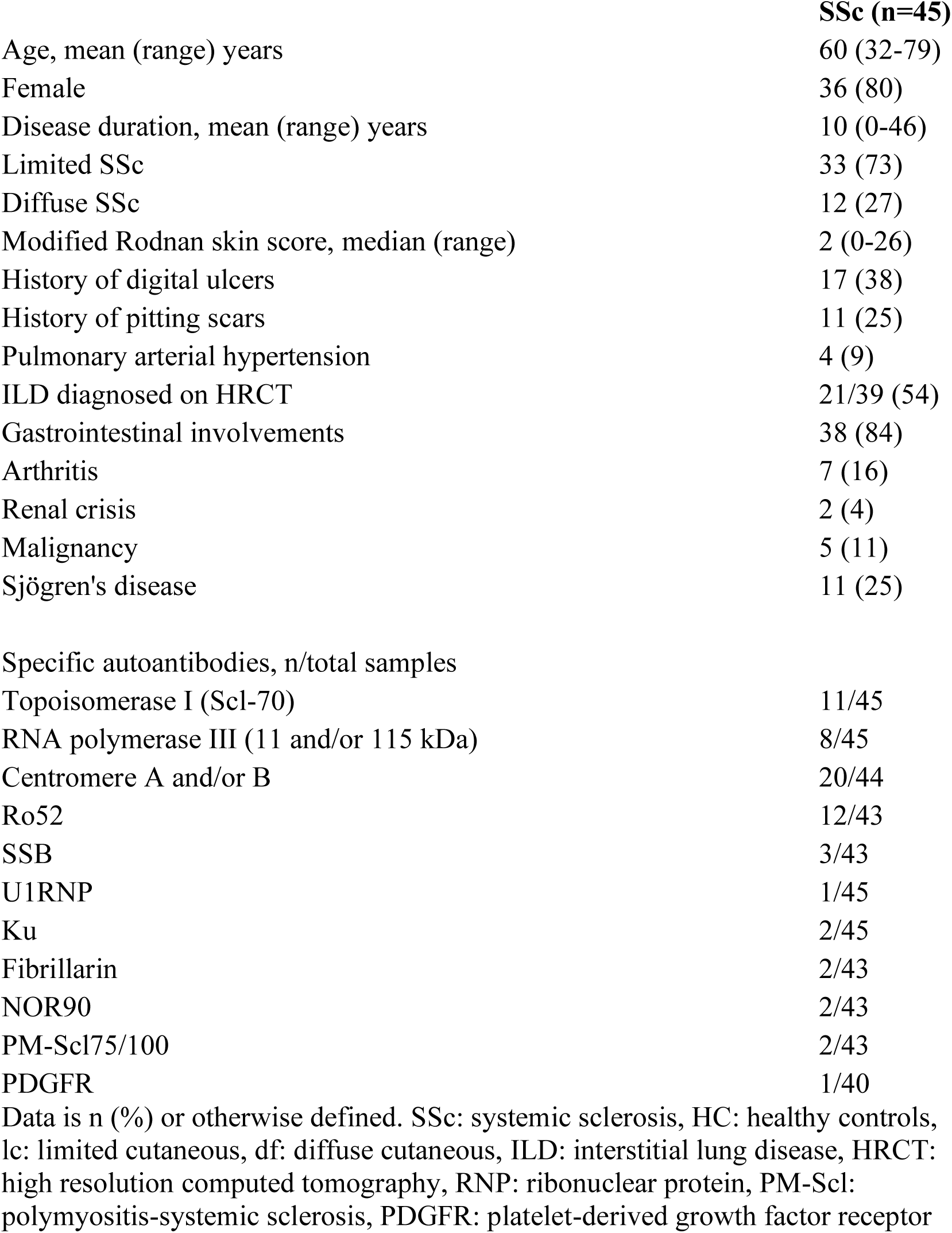
Demographic and clinical characteristics of patients with SSc.

|  | **SSc (n=45)** |
| --- | --- |
| Age, mean (range) years | 60 (32-79) |
| Female | 36 (80) |
| Disease duration, mean (range) years | 10 (0-46) |
| Limited SSc | 33 (73) |
| Diffuse SSc | 12 (27) |
| Modified Rodnan skin score, median (range) | 2 (0-26) |
| History of digital ulcers | 17 (38) |
| History of pitting scars | 11 (25) |
| Pulmonary arterial hypertension | 4 (9) |
| ILD diagnosed on HRCT | 21/39 (54) |
| Gastrointestinal involvements | 38 (84) |
| Arthritis | 7 (16) |
| Renal crisis | 2 (4) |
| Malignancy | 5 (11) |
| Sjögren's disease | 11 (25) |
| Specific autoantibodies, n/total samples |  |
| Topoisomerase I (Scl-70) | 11/45 |
| RNA polymerase III (11 and/or 115 kDa) | 8/45 |
| Centromere A and/or B | 20/44 |
| Ro52 | 12/43 |
| SSB | 3/43 |
| U1RNP | 1/45 |
| Ku | 2/45 |
| Fibrillarin | 2/43 |
| NOR90 | 2/43 |
| PM-Scl75/100 | 2/43 |
| PDGFR | 1/40 |

Data is n (%) or otherwise defined. SSc: systemic sclerosis, HC: healthy controls, lc: limited cutaneous, df: diffuse cutaneous, ILD: interstitial lung disease, HRCT: high resolution computed tomography, RNP: ribonuclear protein, PM-Scl: polymyositis-systemic sclerosis, PDGFR: platelet-derived growth factor receptor

### Cell culture conditions

PBMCs from patients with SSc and HCs were isolated from EDTA-blood by Ficoll-Paque Plus (Cytiva, Malborough, MA, USA) density-gradient centrifugation and separated into a CD14-negative fraction and CD14+ monocytes (≥96% purity) by using CD14 Microbeads and the MACS magnetic separation system (Miltenyi Biotec, Bergisch Gladbach, Germany) (21). The three cell fractions (0.4×10^6^ per well) were incubated with stimulators or medium only in flat-bottomed 96-well cell culture plates for 20 h at 37°C in 5% CO_2_. Cell cultures were set up in duplicates and when indicated supplemented with 500 U/ml IFN-α2b (IntronA; MSD, Rahway, NJ, USA). The number of cell donors used in individual experiments are indicated in text or in the figure legends.

### Cell stimulation with TLR agonists

The cells were stimulated with a TLR7 agonist: RNA containing immune complexes (RNA-IC) consisting of IgG autoantibodies to RNP/Sm and U1snRNP particles, 1 mg/ ml and 2.5 µg/ml, respectively (21, 22). A synthetic TLR8 agonist ORN8L (16, 23) (ChemGenes Corporation, Wilmington, DE, USA) and UV-inactivated herpes simplex virus type I (HSV) sensed by TLR9 (24) were used at concentrations of 0.1 mg/ml and 10% (v/v), respectively (25).

### Detection of type I and III IFNs in cell cultures

Concentrations of IFNs were determined in cell culture supernatants by immunoassays. IFN-α was analyzed by a dissociation-enhanced lanthanide fluoroimmunoassay (DELFIA), that detects most IFN-α subtypes (LLoQ ≥2 U/ml) except IFNα2b (26), used as priming in some cell cultures as indicated.

Concentrations of IFN-β, IFN-λ1 and -λ2 were determined by Bio-Plex Multiplex Cytokine Assay (Bio-Rad, Hercules, CA, USA), with LLoQs of 1, 9 and 10 pg/ml respectively.

Flow cytometric analysis of surface markers and intracellular IFN-α, and autoantibody analyses are described in the Supplementary method file.

## STATISTICS

Continuous variables are presented as medians with interquartile ranges (IQR). IFN values were log-transformed when appropriate. Groups were compared using Mann-Whitney U-test, Wilcoxon signed-rank test, Kruskal Wallis or Friedman’s tests as appropriate, using SPSS software (29.0.2). P-values ≤ 0.05 were considered significant.

## RESULTS

### TLR agonist-mediated induction of IFN-α

We investigated whether patients with SSc responded to TLR agonists (RNA-IC, ORN8L, and HSV) by producing IFN-α, and if their IFN-α producing capacity differs from that of HCs. All three stimuli significantly induced IFN-α production in both total PBMCs and CD14-negative (monocyte-depleted) PBMCs (Fig. 1A–B). However, IFN-α levels were consistently lower in SSc PBMCs compared to HCs (p< 0.001-0.02; Fig. 1A).

**Figure 1.**
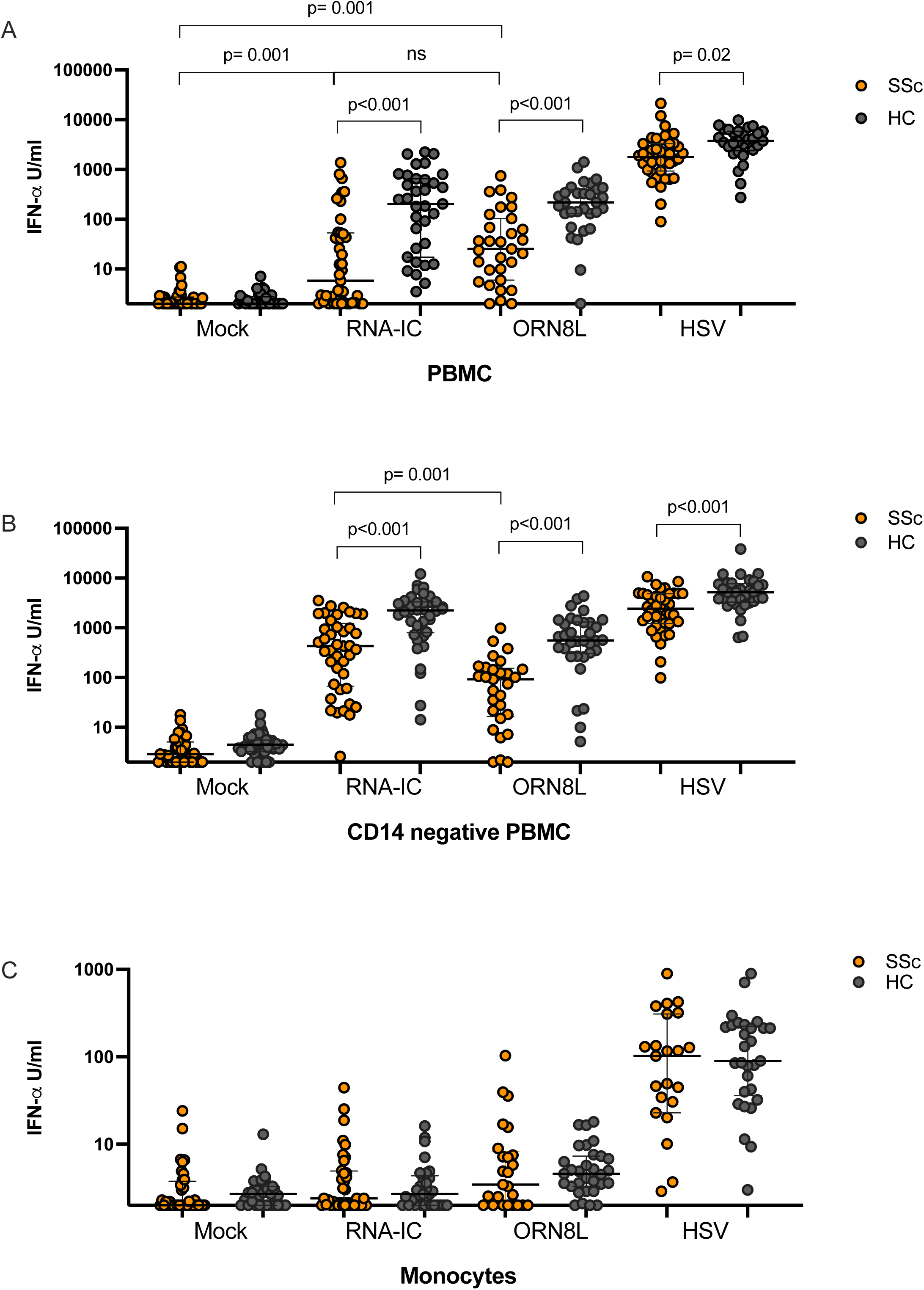
(A-C) IFN-α production in cell cultures with peripheral blood mononuclear cells (PBMC) and PBMC depleted of monocytes (CD14 negative PBMC) and monocytes, stimulated with (A) RNA-IC containing immune complexes (RNA-IC), (B) a TLR8 agonist ORN8L or (C) inactivated herpes simplex virus type I (HSV) from patients with systemic sclerosis (SSc, orange dots) and healthy controls (black dots). IFN-α (U/ml) in the cell culture supernatants were analyzed after 20h with an immunoassay detecting most IFN-α subtypes. Differences between groups were analyzed with Mann Whitney U test or Kruskal Wallis test and considered significant if p≤0.05. Every dot corresponds to one unique cell donor. The plots show median (horizontal bars) with interquartile range.

Despite lower levels, most SSc patients produced detectable IFN-α: 77% (33/43) responded to RNA-IC (median: 9.4 U/ml [IQR: 98.5]) and 93% (29/31) to ORN8L (median: 22 U/ml [IQR: 86]). HSV induced markedly higher IFN-α than RNA-IC (median: 1770 U/ml [IQR: 2325]) or ORN8L (both p < 0.01). Mock stimulation yielded low or no IFN-α, with no differences between groups.

To delineate the contribution of monocytes, IFN-α production was measured in CD14-depleted PBMCs. RNA-IC triggered >70-fold higher IFN-α levels in CD14-negative PBMCs (median: 438 U/ml [IQR: 1321], n=39) compared to total SSc PBMCs. ORN8L induced a modest increase (median: 77 U/ml [IQR: 123], p=0.23), while HSV-induced IFN-α was unaffected by monocyte depletion. Lower IFN-α levels consisted in SSc PBMCs compared to HCs also after monocyte depletion, irrespective of stimulus (Fig. 1B).

Monocytes produced little IFN-α in response to RNA-IC (median: 3 U/ml [IQR: 2.6]) or ORN8L (median: 4.4 U/ml [IQR: 4.3]; Fig.1C), whereas HSV triggered modestly significant levels (SSc; median 116 U/ml [IQR: 290]). IFN-α production by monocytes did not differ between SSc patients and HCs for any stimulus applied (Fig. 1C). In summary, SSc PBMCs produced variable but lower IFN-α levels than HCs in response to TLR7/8/9 stimuli.

### IFN-β production across cell subsets

Given the variability in IFN-α production, we investigated the IFN-β producing capacity of blood leukocytes in SSc. IFN-β levels were measured in PBMCs, CD14-negative PBMCs and monocytes from SSc patients and HCs. RNA-IC stimulation induced highly variable IFN-β responses in PBMCs (SSc: median 2.0 pg/ml, range: 444, n=26; HC: 14 pg/ml, range 636, n=22). Moreover, RNA-IC-induced IFN-β levels increased significantly upon monocyte depletion (SSc: 52 pg/ml [IQR:116]; HC: 81 pg/ml [IQR:147], both p<0.001; Fig. 2A-B). In contrast, ORN8L and HSV induced IFN-β across all cell fractions, irrespective of monocyte presence (Fig. 2A-C). In contrast to IFN-α, there was no difference in IFN-β production between SSc patients and HCs, except for lower response among ORN8L-stimulated CD14-negative PBMCs from SSc patients (Fig. 2B), and higher IFN-β production in monocytes from SSc patients compared to HCs (SSc: 342 pg/ml [IQR:1491] vs. HC pg/ml 59.9 [IQR:364], p=0.041; Fig. 2C). These findings indicate that ORN8L and HSV induce strong IFN-β production in both PBMC and monocytes, whereas RNA-IC-induced response in PBMC is attenuated by monocytes.

**Figure 2.**
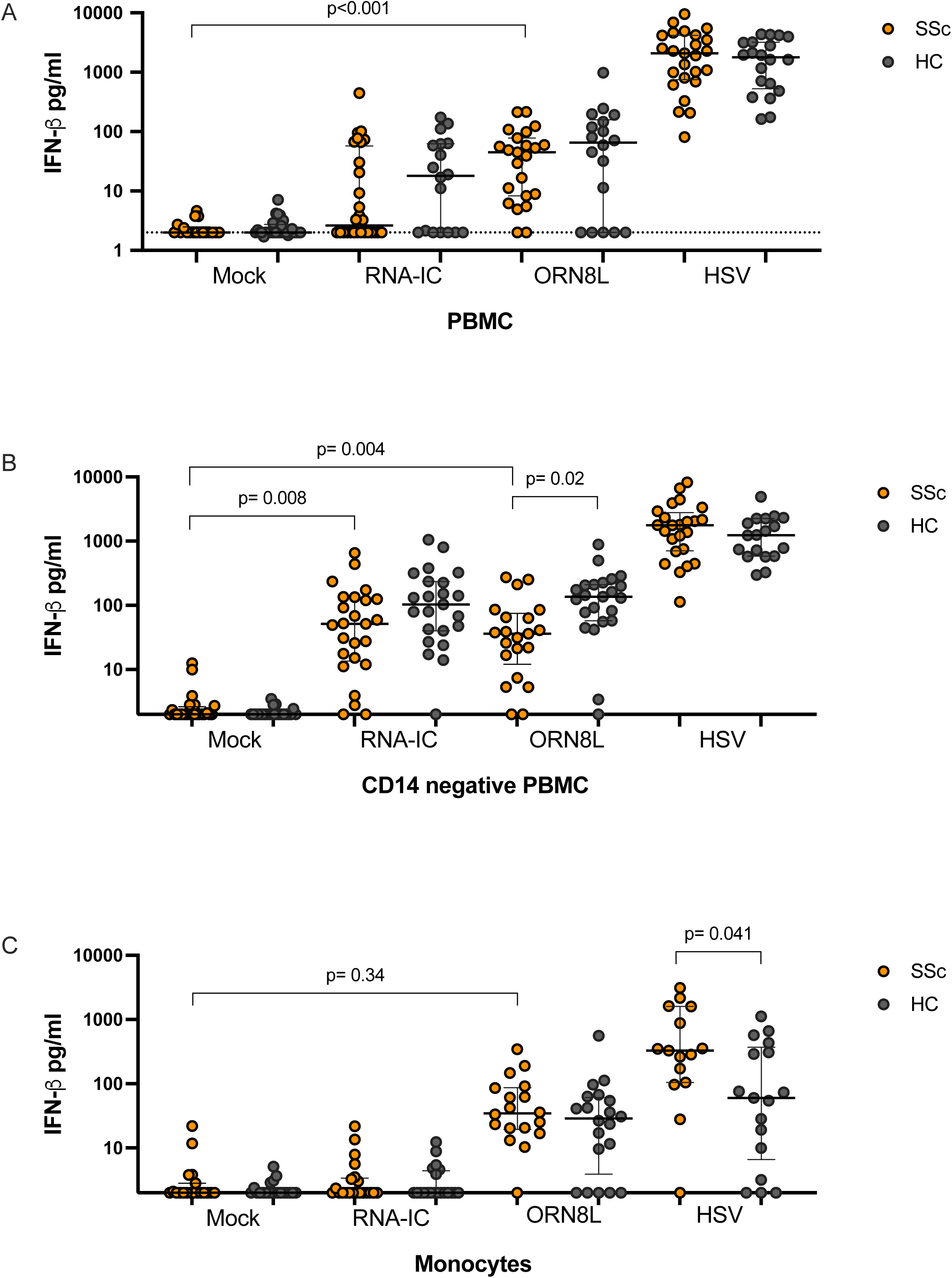
(A-C) IFN-β production in cell cultures with peripheral blood mononuclear cells (PBMC) and PBMC depleted of monocytes (CD14 negative PBMC) and monocytes, stimulated with (A) RNA-IC containing immune complexes (RNA-IC), (B) a TLR8 agonist ORN8L or (C) inactivated herpes simplex virus type I (HSV) from patients with systemic sclerosis (SSc, orange dots) and healthy controls (black dots). Concentration of IFN-β (pg/ml) was analyzed by a bead-based immunoassay. Differences between groups were analyzed with Mann Whitney U test or Kruskal Wallis test and considered significant if p≤0.05. Every dot corresponds to one unique cell donor. The plots show median (horizontal bars) with interquartile range.

### IFN-λ2 and -λ1 production by SSc blood leukocytes

IFN-λs primarily act against pathogens but also have other immunomodulatory functions (27), though less studied than type I IFNs. Therefore, we assessed IFN-λ1 and -λ2 responses to the TLR agonists RNA-IC, ORN8L and HSV by PBMC, CD14-negative PBMC and monocytes from SSc patients and HCs.

All three stimuli induced IFN-λ2 in PBMC and CD14-negative PBMC (Fig. 3A-B), whereas only ORN8L and HSV triggered significant production in monocytes (Fig. 3C). Monocyte depletion increased RNA-IC-induced IFN-λ2 in both SSc and HCs (p=0.002), whereas only HC PBMC (p=0.008) increased their IFN-λ2 production in response to ORN8L (Fig.3A–3B).

**Figure 3.**
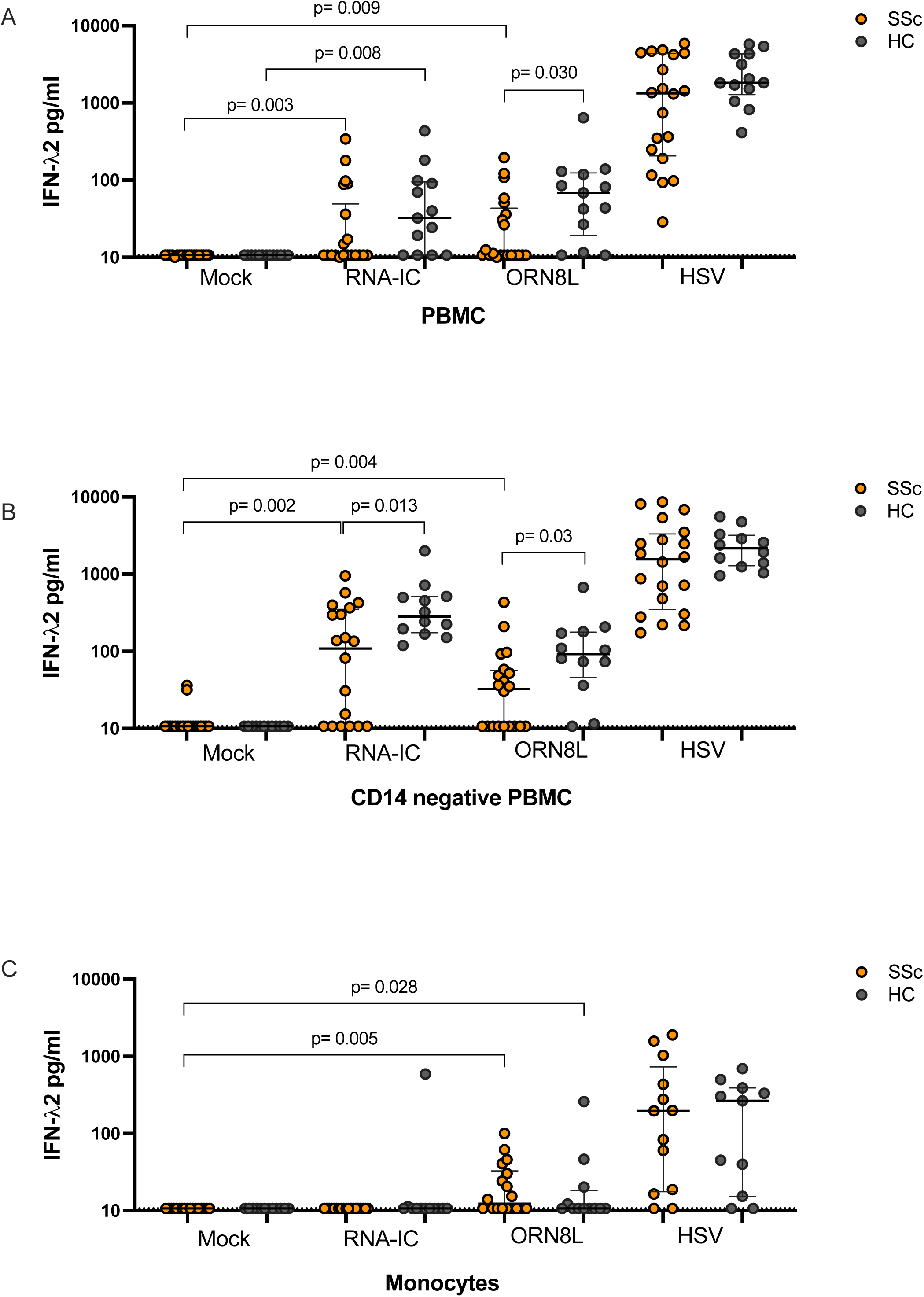
(A-C) IFN-λ2 production in cell cultures with peripheral blood mononuclear cells (PBMC) and PBMC depleted of monocytes (CD14 negative PBMC) and monocytes, stimulated with (A) RNA-IC containing immune complexes (RNA-IC), (B) a TLR8 agonist ORN8L or (C) inactivated herpes simplex virus type I (HSV) from patients with systemic sclerosis (SSc, orange dots) and healthy controls (black dots). Concentration of IFN-λ2 (pg/ml) was analyzed by a bead-based immunoassay. Differences between median values were analyzed with Mann Whitney U test or Kruskal Wallis test and considered significant if p≤0.05. Every dot corresponds to one unique cell donor. The plots show median (horizontal bars) with interquartile range.

Across all conditions, IFN-λ2 production was 5-20-fold higher than IFN-λ1 in both PBMC and CD14-negative PBMC cell cultures (Fig. 3A-B and supplementary fig. 1A-B). Only HSV induced IFN-λ1 in monocytes with no difference between SSc and HCs (supplementary fig 1C).

Monocyte depletion slightly increased FN-λ1 in both SSc (p=0.017) and HCs (p=0.028). Notably, the proportion of SSc responding to RNA-IC increased from 55% to 81% after monocyte depletion (supplementary Fig. 1B). In contrast, monocyte depletion did not enhance IFN-λ1 responses to ORN8L.

In summary, IFN-λ2 was produced at higher levels than IFN-λ1 across most conditions. IFN-λ2 production was generally higher in HC cultures than in SSc, except in response to HSV where levels were comparable. Monocyte depletion significantly increased RNA-IC-induced IFN-λ1 and IFN-λ2 levels and neither cytokine was produced by monocytes in response to RNA-IC.

### Effect of IFN-α priming on IFN production

We previously demonstrated that IFN-α2b-priming enhances IFN-α production in HC PBMCs and pDCs (18, 29). Herein, we found that priming of SSc PBMCs substantially increased IFN-α production in response to RNA-IC (from 7.6 to 74 U/ml, n=42, p<0.001) and ORN8L (from 28 to 108 U/ml, n=31, p<0.001), but not in HSV-stimulated PBMC cultures (supplementary fig. 2A). IFN-β and IFN-λ2 also increased modestly following IFN-α2 priming during RNA-IC and ORN8L stimulation (supplementary Figs. 2B-2C).

### Proportion of plasmacytoid dendritic cells and monocytes, and identification of IFN-α producing cells

To determine whether differences in IFN-α production between SSc patients and HCs were due to altered proportions of pDCs and monocytes in SSc, we performed flow cytometric analysis of PBMCs. There was no difference in median percentage of CD14+ monocytes between SSc patients (23%, IQR: 10.6, n=40) and HCs (20%, IQR: 13.6, n=19, p=0.118). In contrast, SSc patients had a lower percentage of pDCs (0.24%, IQR: 0.17) compared to HCs (0.44%, IQR: 0.21, p< 0.001).

To identify the cellular source of IFN-α, we stained PBMCs for intracellular IFN-α and surface markers specific for pDCs (BDCA2) and monocytes (CD14), following stimulation with RNA-IC, ORN8L or HSV.

The highest percentage of IFN-α-producing cells was observed after HSV stimulation. Intracellular IFN-α was detected exclusively within the pDC population, accounting for 8.1% of all pDCs in SSc PBMCs (range: 5.6-10.9%, SD: 2.2%, n=4; supplementary Fig. 3A). This proportion was markedly higher in HCs (22± 4.7%, n=2) (supplementary table 2). No IFN-α positive cells were detected with an isotype IgG or in mock-stimulated PBMCs (Supplementary fig. 3B-C) or in CD14+ monocytes among PBMCs (Supplementary fig. 3 D). In response to RNA-IC and ORN8L, <1% of pDCs from SSc patients produced IFN-α, compared to ≈3.8% in HCs (supplementary table 2).

In summary, HSV induced significantly higher proportion of IFN-α-producing pDCs in SSc PBMC compared to RNA-IC and ORN8L, although the proportion was lower than in HCs.

### Associations between IFN production and clinical manifestations in patients with SSc

Next, we investigated whether type I IFN triggered by RNA-IC, ORN8L or HSV, was associated with specific clinical characteristics in SSc patients.

Comparing IFN-α levels in the lcSSc and dcSSc subtypes we found that patients with dcSSc produced higher levels of IFN-α in response to HSV (median: 3379 vs. 1543 U/ml; p= 0.037; Fig. 4A). No subtype differences were observed for IFN-β, IFN-λ1 or -λ2 (data not shown).

**Figure 4.**
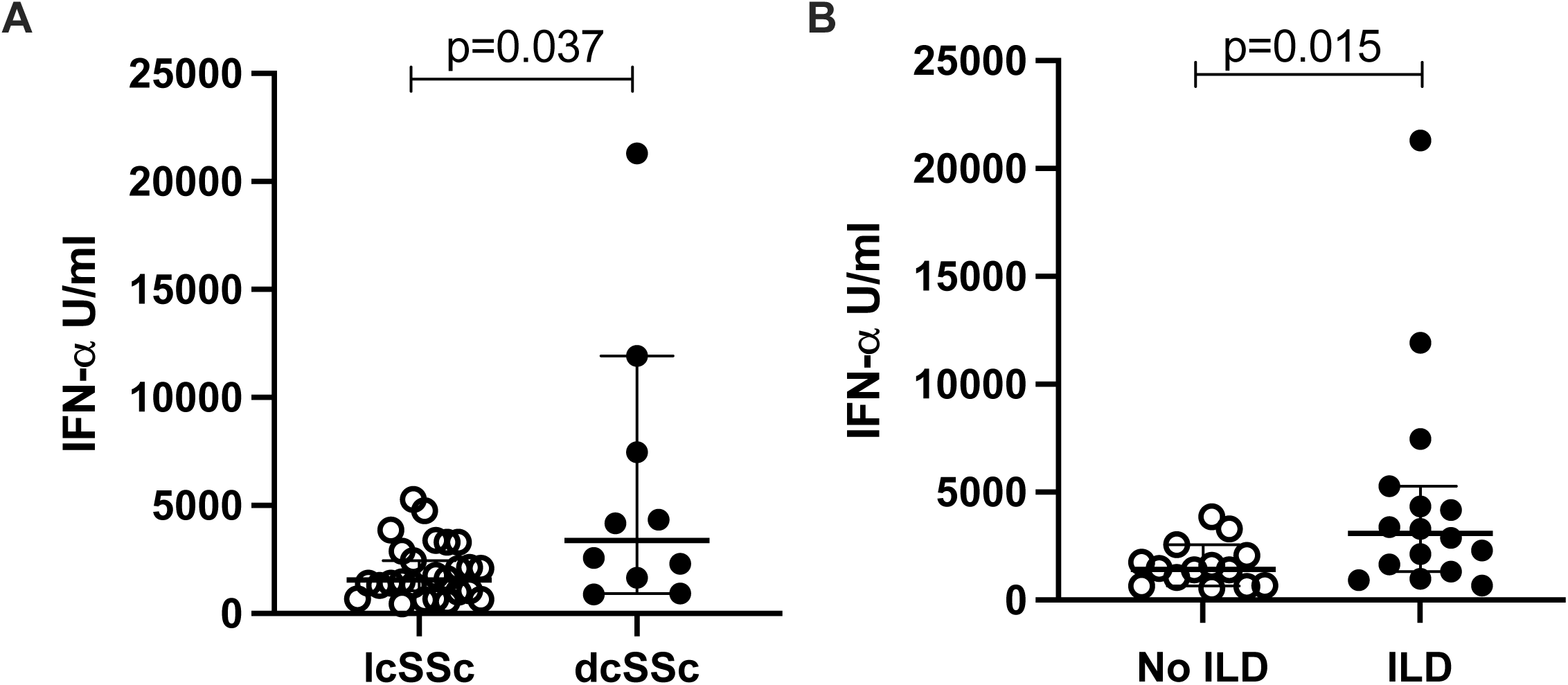
IFN-α production by PBMC isolated from patients with systemic sclerosis (SSc) with limited cutaneous (lcSSc) or diffuse cutaneous (dcSSC) subtype and (B) without or with interstitial lung disease (ILD). Peripheral mononuclear cells (PBMC) were stimulated with inactivated herpes simplex virus type I (HSV) for 20h. IFN-α (U/ml) was analyzed in cell culture supernatants by an immunoassay recognizing most IFN-α subtypes. Differences between median values were analyzed with Mann Whitney U test and considered significant if p≤0.05. The plots show median (horizontal bars) with interquartile range.

SSc patients with ILD on HRCT had higher HSV-induced IFN-α levels (median: 3098 U/ml, IQR=3635, n=16) compared to patients without ILD (1435 U/ml, IQR:1530, n=14, p=0.015; Fig. 4B). No association was found between IFN-β levels and ILD (data not shown).

Calcium channel blocker (CCB), especially nifedipine, treatment was associated with reduced IFN-α levels in PBMCs in response to HSV (n=33, p=0.021) (supplementary Fig. 4) and RNA-IC (p=0.036, not shown).

No significant associations were found between induced IFN levels and occurrence of PAH, digital ulcers, pitting scars, cancer, Sjögren’s disease, methotrexate treatment or the modified Rodnan skin score.

### Association between SSc specific autoantibodies and IFN production

SSc-specific autoantibodies were detected in 43 of 45 of patients (Table 1). We examined whether autoantibodies to RNApol III, topoisomerase I or centromere proteins were associated with IFN-α production in response to RNA-IC, ORN8L or HSV. Furthermore, IFN-α levels were compared between patients with medium-to-strong antigen reactivity, and those lacking the specificity.

Patients with strong anti-RNApol III (155/11 kDa) reactivity showed a significantly higher IFN-α production by PBMC in response to HSV (median: 4187 vs. 1638 U/ml, p=0.011), n= 4/36; Fig. 5A). No difference was found for anti-topoisomerase I (median: 1653 vs. 1904 U/ml, n=8/36, p=0.68; Fig. 5B) while strong CENP-B/A reactivity was associated with lower HSV-induced IFN-α levels (median: 1401 vs. 2444 U/ml, n= 17/39, p=0.005; Fig. 5C). Similar trends were seen in CD14-negative PBMC. No association to other autoantibodies was detected (data not shown).

**Figure 5.**
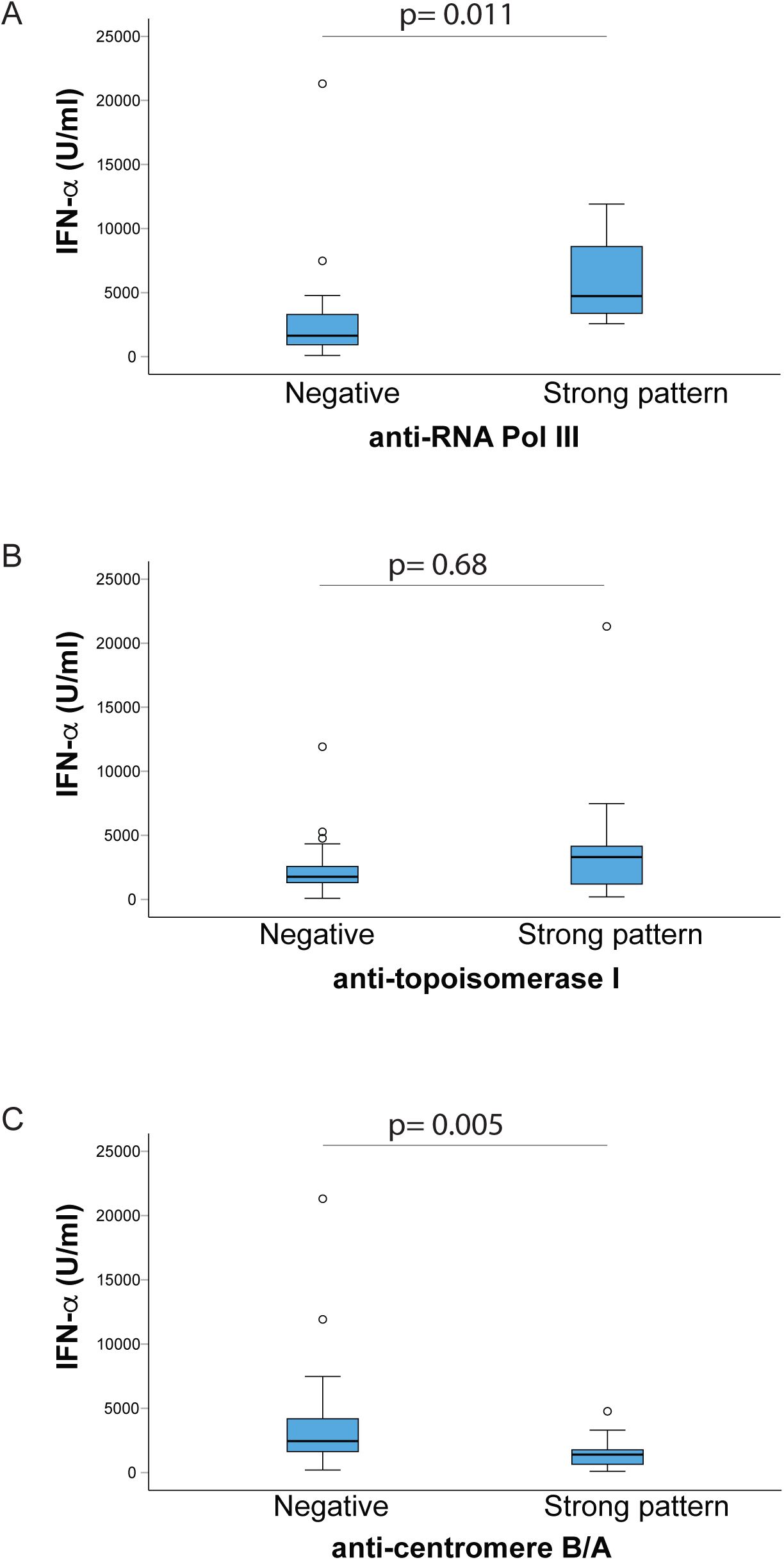
IFN-α production stratified by autoantibody specificities in patients with systemic sclerosis. The patients were divided regarding presence of strong reactivity to (A) RNA polymerase III, topoisomerase I or (C) centromere versus lack of the specific autoantibody as determined with Line Immuno Assay (LIA). Peripheral mononuclear cells (PBMC) were stimulated with inactivated herpes simplex virus type I (HSV) for 20h. IFN-α (U/ml) was analyzed in cell culture supernatants by an immunoassay recognizing most IFN-α subtypes. Differences between groups were analyzed with Mann Whitney U test and considered significant if p≤0.05. The box plots show median (horizontal bars) with interquartile range, minimal and maximal values.

## DISCUSSION

Our study demonstrates that PBMCs from patients with SSc have the capacity to produce type I and III IFNs in response to TLR7/8/9 stimulation with varying influence of monocytes, and the viral stimulus being the most potent IFN inducer.

Nucleic acid-containing immune complexes have been studied extensively in SLE and are implicated as central drivers of the chronically activated type I IFN system (28). Such endogenous stimuli could also be present in SSc, continuously activating IFN production. Herein we showed that RNA-IC induced variable but lower levels of IFNs in SSc PBMCs than in HCs. However, priming with IFN-α markedly enhanced its own production when triggered by RNA-IC or ORN8L, and increased the IFN-β and IFN-λ2 levels although more modest. Such amplification may increase the total IFN exposure of various target organs in SSc.

We found that the TLR8 agonist ORN8L induced IFN-α production in CD14-negative PBMCs, supporting previous reports of TLR8-mediated IFN-α expression in SSc pDCs in response to ORN8L (16).

These findings contrast with a recent study reporting the absence of TLR8 in SSc pDCs and a lack of IFN-α production upon TLR8 stimulation (29). This discrepancy may reflect differences in agonist structure, experimental methodology, or contributions from additional cell types beyond pDCs and monocytes.

Notably, monocyte depletion did not affect IFN responses to ORN8L, in contrast to RNA-IC induced IFN, which was increased in both SSc and HCs. This suggests a potential regulatory role for monocytes in TLR7-mediated type I and III IFN production, consistent with prior findings for IFN-α in HCs (15). Furthermore, we observed that ORN8L robustly induces IFN-β and IFN-λ2 production in SSc monocytes, a cell type known to express TLR8 (16). Collectively, our data support a role for TLR8-driven IFN expression in SSc involving both monocytes and pDCs.

Several viruses have been investigated as triggering factors for SSc, though no direct causal relationship has been established (30). In our study, HSV induced significantly higher levels of IFNs than RNA-IC or ORN8L and stimulated comparable levels of IFN-α and IFN-λ1/2 in monocyte cultures from both SSc patients and HCs. These findings are in line with previous reports identifying HSV as a strong IFN-α inducer in PBMCs from both HCs and patients with SLE (31). Interestingly, HSV-induced IFN-β production was even higher in SSc monocytes compared to HCs. While some studies have reported increased proportion of monocytes in SSc, others, including our own, did not observe significant differences (32). These latter findings suggest that the elevated IFN-β production by SSc monocytes, compared to HCs, is not attributable to increased monocyte numbers, but might reflect intrinsic functional alterations. One possible explanation for the high HSV-induced IFN production could be an upregulated TLR9 expression in SSc leukocytes (32).

Thus, an indirect but important contribution of viruses to SSc pathogenesis could be mediated through a strong IFN response, acting as in vivo priming and amplifying the IFN-inducing capacity of endogenous IFN-inducers such as nucleic-acid-containing ICs.

Overall, the type I and III IFN production by monocytes in response to HSV underscores their potential pathological role in SSc, especially considering their higher abundance in blood and tissues compared to pDCs, as well as their implicated role in fibrosis (33).

pDCs are considered as professional IFN-α producers, but with certain stimuli monocytes can also be triggered to type I IFN production. Our flow cytometric analysis of PBMCs revealed pDCs, but not CD14+ monocytes, as the main cellular source of IFN-α, regardless of stimulus. In contrast, IFN-α levels were elevated in HSV-stimulated monocyte cultures. Given the >90% purity of the monocyte preparations, pDC contamination is unlikely to explain this finding. Instead, it probably reflects low per-cell IFN-α production by monocytes, in contrast to the high-output profile of pDCs (31). These findings support the notion that monocytes, although less potent on a per-cell basis, can contribute to the overall interferon response in SSc. Regarding the pDCs in SSc, we find less than 1% to produce IFN-α when stimulated with RNA-IC or ORN8L, mirroring the levels in the stimulated cell cultures. These findings confirm the previous observations in HC, demonstrating that only a small subset of activated pDCs produce IFN-α (21) and IFN-λ (18). Whether this reflects functionally different pDC subsets or temporal activation as described for other virus induced genes remain to be determined (34).

Finally, we investigated a potential association between IFN levels, clinical manifestations, and the presence of three major SSc-specific autoantibodies. Interestingly, elevated IFN-α production was significantly associated with dcSSc and ILD. Our findings align with prior reports linking elevated serum IFN scores, based on IFN-regulated chemokines, to poor pulmonary outcome (4, 10). They also corresponded with increased migration of pDCs to lungs and skin in SSc, along with reduced numbers in circulation (35). Additionally, a recent multi-omic study demonstrated an upregulated of IFN gene signature in SSc patients with ILD compared to those without ILD (36).

Unexpectedly, IFN-α but not IFN-β or IFN-λ-levels, were reduced in stimulated PBMC cultures from patients treated with CCBs, particularly nifedipine, the most frequently used CCB in our study population. To our knowledge, this is the first report of such an association for nifedipine, although a previous study described its inhibition of IFN-γ production in PBMCs in vitro (37). Another CCB has also been shown to reduce TNF-α and MCP-1 in activated PBMCs (38). Whether a similar mechanism underlies suppression of IFN-α by CCBs remains to be explored.

We did not observe an association between IFN levels and anti-topoisomerase I antibodies and, despite the known link between anti-topoisomerase I and ILD (3). This finding aligns with our previous work showing no association between sera from anti-topoisomerase I positive SSc patients and IFN-α production by healthy PBMC (12). Interestingly, induced IFN-α levels were significantly elevated in patients with a strong anti-RNApol III reactivity, while patients with anti-CENP-A or -B antibodies exhibited lower IFN-α production. Our results are consistent with the clinical observations that anti-CENP antibodies correlate with milder disease, whereas anti-RNApol III antibodies are linked to dcSSc, rapid progression of skin thickening, SSc renal crisis, and malignancy (39). We found no significant association with renal crisis or malignancy, possibly due to their low prevalence in our study.

Our study has several strengths. Patients were well characterized by specialist rheumatologists at a single clinic, minimizing the variability in clinical assessment. Importantly, we directly measured the IFN producing capacity of freshly isolated patient and controls cells, rather than relying on indirect methods such as analysis of IFN stimulated genes/proteins. Our study also has limitations. First, the number of patients with certain clinical phenotypes was too small to detect potential associations with IFN levels. Second, not all cell fractions from each patient were stimulated with all three TLR agonists, which may have limited our ability for more comprehensive comparisons across conditions.

We conclude that viral stimuli trigger high levels of both type I and III IFNs by different cell types and could prime IFN production by less powerful endogenous stimuli such as self-nucleic-containing immune complexes. A perpetuated type I and III IFN production via multiple pathways may fuel both local and systemic immune dysregulation contributing to pathogenesis and specific manifestations in SSc, such as ILD.

## Data Availability

All data produced in the present work are contained in the manuscript

## ACKNOWLEDGEMENT

We thank Neha Behare for excellent technical assistance and research nurse Rezvan Kiani Dehkordi for collecting the blood samples.

## Notes

### Competing Interest Statement

The authors have declared no competing interest.

### Funding Statement

The work was supported by grants from the Swedish Rheumatism Association, the Uppsala University Hospital Development Foundation; Department of Medical Sciences, Uppsala University, Sweden., the King Gustaf V 80-year Foundation.

### Author Declarations

Ethics committee of Swedish National Etikprovningsmyndigheten gave ethical approval for this work.

