## Supplementary Material for "Induction of type I and III interferons by viral and endogenous stimuli in systemic sclerosis"

### Supplementary files

| **Supplementary Table 1**<br>**Medication of SSc patients** | |
|---|---|
| | **SSc (n=45)** |
| | n (%) |
| Calcium channel blockers | 32 (71) |
| Nifedipine | 27 (60) |
| Bosentan | 5 (11) |
| Iloprost | 4 (9) |
| Phosphodiesterase 5 inhibitors (sildenafil) | 9 (20) |
| Prednisolone | 4 (9) |
| Methotrexate | 7 (16) |
| Hydroxychloroquiene | 3 (7) |
| Secukinumab | 1 (2) |
| Azathioprine | 1 (2) |
| Mycophenolate mofetil (MMF) | 5 (11) |
| Cyclophosphamide | 1 (2) |
| Tozilizumb | 2 (4) |
| Rituximab | 1 (2) |
| Nintedanib | 2 (4) |

**Supplementary Table 2.** Detection of IFN-α producing pDCs by flow cytometric staining after stimulation with herpes simplex virus type I

| Cell donor | pDC/PBMC % | IFN-α+ pDC % RNA-IC | IFN-α+ pDC % ORN8L | IFN-α+ pDC % HSV |
|---|---|---|---|---|
| HC1 | 0.51 | 1.32 | 0.96 | 18.7 |
| HC2 | 0.62 | 6.21 | 6.67 | 25.4 |
| *Mean ±SD* | ***0.57±0.08*** | ***3.8±3.5*** | ***3.8±4.1*** | ***22.1±4.7*** |
| SSc1 | 0.12 | 0.6 | 1.11 | 7.51 |
| SSc2 | 0.27 | 0 | 0.75 | 8.5 |
| SSc3 | 0.33 | 0.25 | 0.50 | 5.62 |
| SSc4 | 0.50 | 0.32 | 0.70 | 10.9 |
| *Mean ±SD* | ***0.31±0.16*** | ***0.29±0.25*** | ***0.77±0.25*** | ***8.1±2.2*** |

### Supplementary figures

Suppl. figure 1

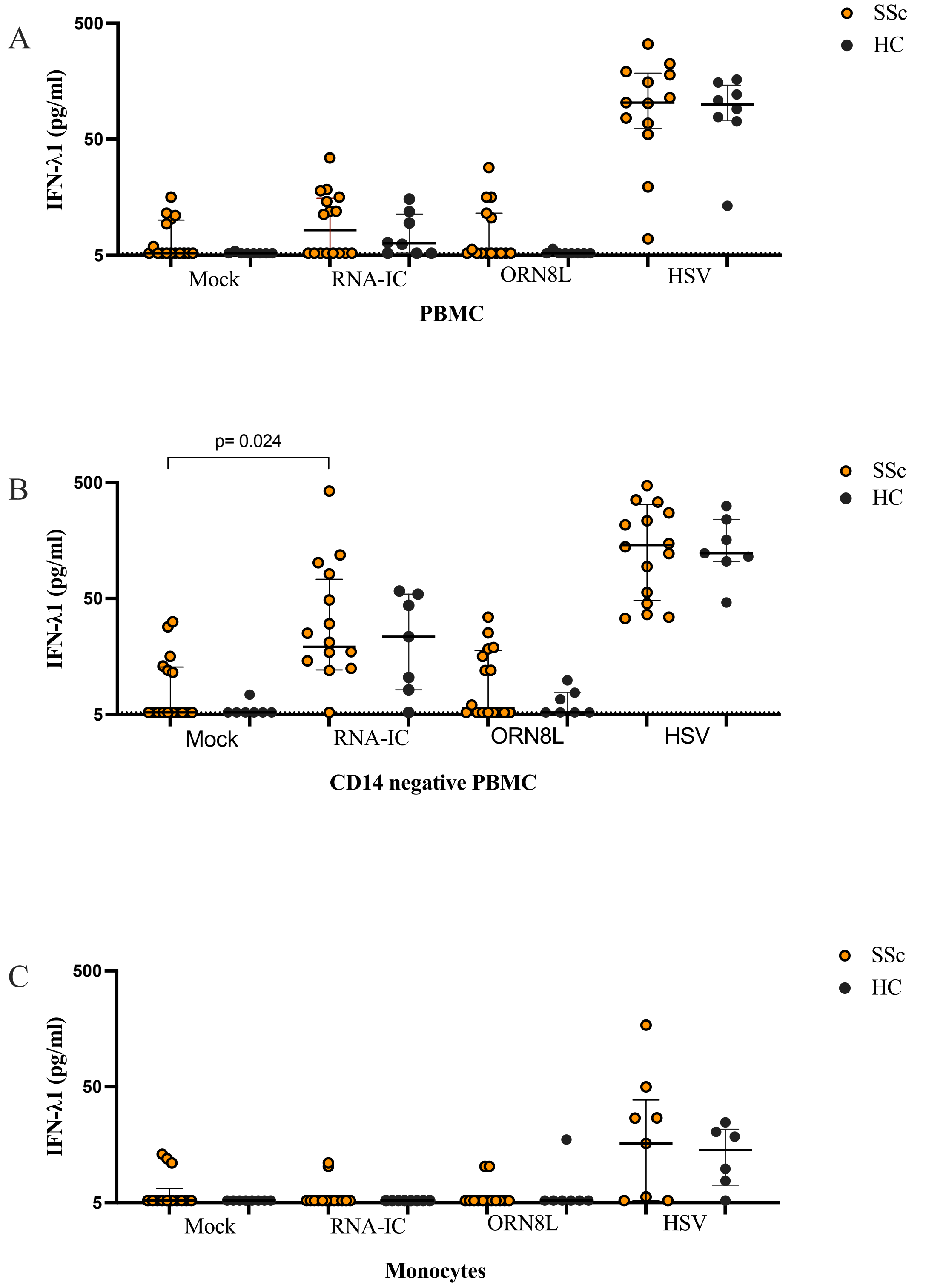

**Supplementary figure 1**

(A-C) IFN-λ1 production in cell cultures with peripheral blood mononuclear cells (PBMC) and PBMC depleted of monocytes (CD14 negative PBMC) and monocytes, stimulated with (A) RNA-IC containing immune complexes (RNA-IC), (B) a TLR8 agonist ORN8L or (C) inactivated herpes simplex virus type I (HSV) from patients with systemic sclerosis (SSc, orange dots) and healthy controls (black dots). Concentration of IFN-λ1 was analyzed by a bead-based immunoassay. Differences between groups were analyzed with Mann Whitney U test or Kruskal Wallis test and considered significant if $p \leq 0.05$. Every dot corresponds to one unique cell donor. The plots show median (horizontal bars) with interquartile range.

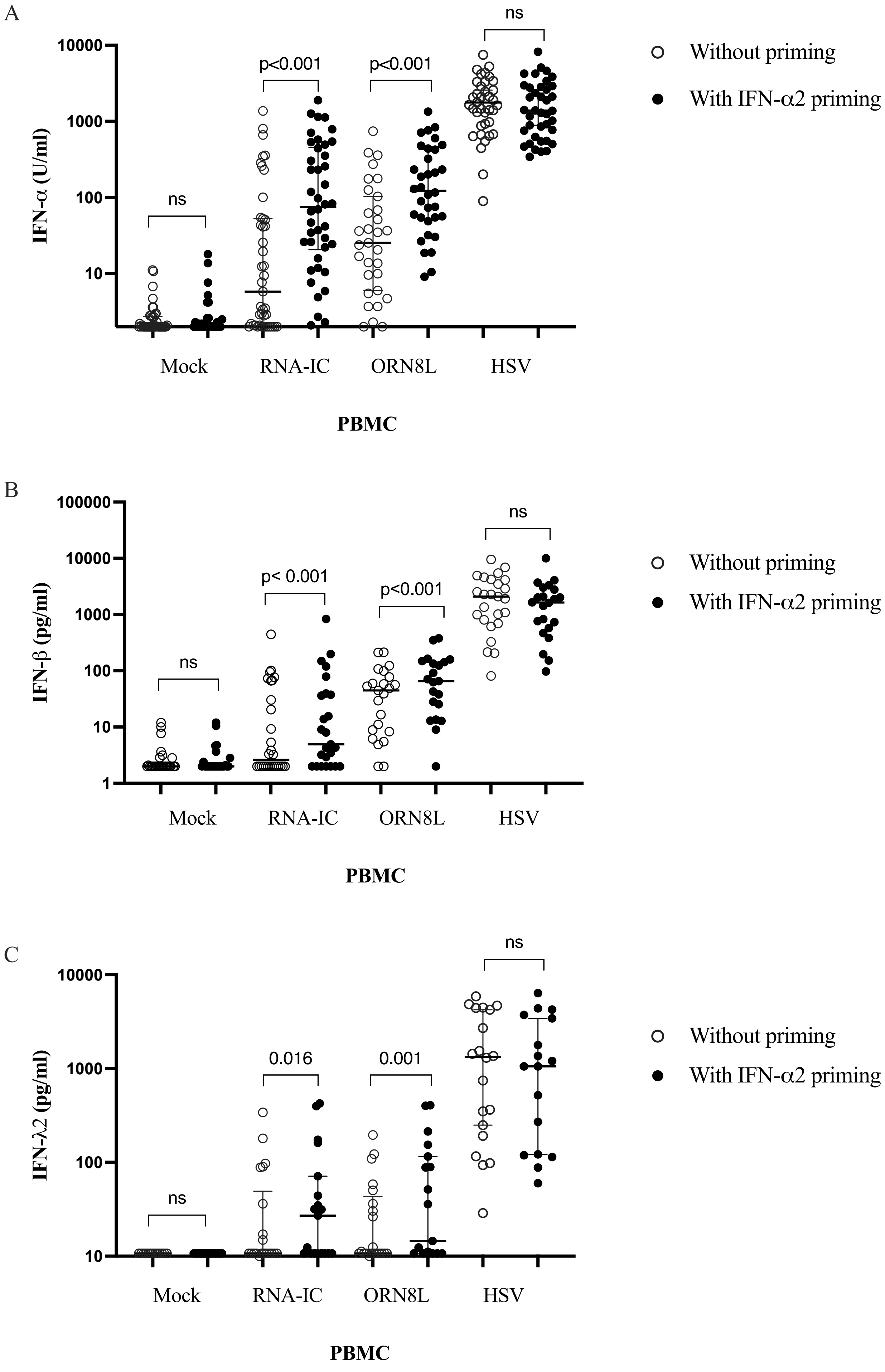

A
IFN-α (U/ml)
10000
1000
100
10
ns
p<0.001
p<0.001
ns
Mock
RNA-IC
ORN8L
HSV
PBMC
Without priming
With IFN-α2 priming
B
IFN-β (pg/ml)
100000
10000
1000
100
10
1
ns
p< 0.001
p<0.001
ns
Mock
RNA-IC
ORN8L
HSV
PBMC
Without priming
With IFN-α2 priming
C
IFN-λ2 (pg/ml)
10000
1000
100
10
ns
0.016
0.001
ns
Mock
RNA-IC
ORN8L
HSV
PBMC
Without priming
With IFN-α2 priming

**Supplementary figure 2**

Effect of IFN-α2b "priming" on (A) IFN-α, (B) IFN-β and (C) IFN-λ2 production by peripheral mononuclear cells (PBMC) from patients with systemic sclerosis were stimulated with medium only (Mock), RNA-IC containing immune complexes (RNA-IC), a TLR8 agonist ORN8L or inactivated herpes simplex virus type I (HSV), in presence or absence of IFN-α2b ("priming") for 20h. The cell culture supernatants were analyzed after 20h with an immunoassay detecting most IFN-α subtypes but not IFN-α2b used for priming. IFN-β and IFN-λ2 concentrations were analyzed by a bead-based immunoassay. Differences between groups were analyzed by Kruskal Wallis test and considered significant if $p \leq 0.05$. Every dot corresponds to one unique cell donor. The plots show median (horizontal bars) with interquartile range.

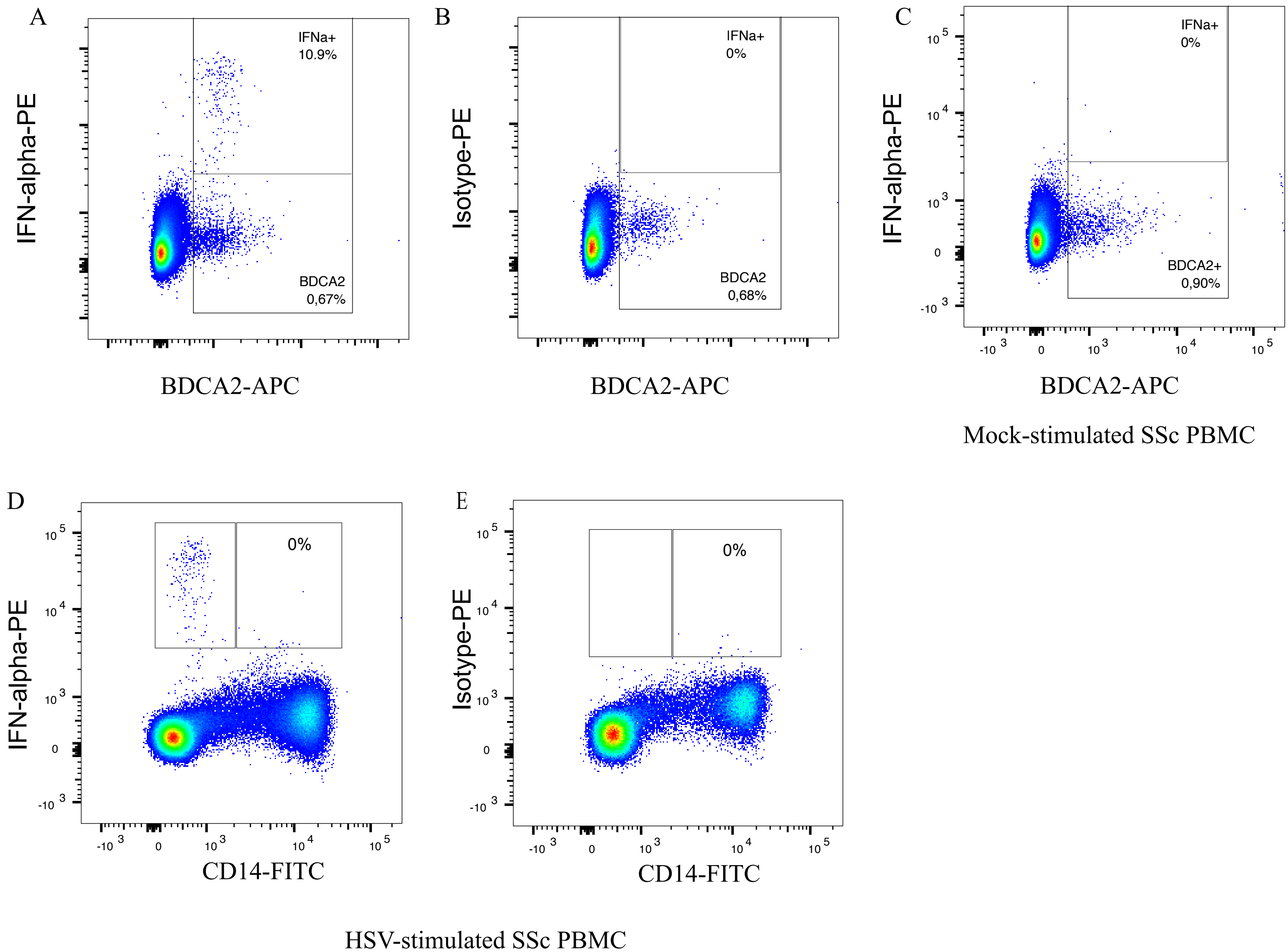
A
IFN-alpha-PE
IFNa+
10.9%
BDCA2
0,67%
BDCA2-APC
B
Isotype-PE
IFNa+
0%
BDCA2
0,68%
BDCA2-APC
C
IFN-alpha-PE
IFNa+
0%
BDCA2+
0,90%
BDCA2-APC
Mock-stimulated SSc PBMC
D
IFN-alpha-PE
0%
CD14-FITC
E
Isotype-PE
0%
CD14-FITC
HSV-stimulated SSc PBMC

**Supplementary figure 3**

Identification of IFN-α producing cells by flow cytometric analysis among peripheral blood mononuclear cells (PBMC) from patients with systemic sclerosis after stimulation with (A, B, D and E) inactivated herpes simplex virus type I (HSV) or (C) medium only (Mock) for 9 hours, the last three hours in presence of Brefeldin A. (A, C, D) The cells were stained with fluorochrome labelled monoclonal antibodies to intracellular IFN-α and (A, B, C) anti-BDCA2 antibody identifying plasmacytoid dendritic cells (pDCs) or by using (B, E) isotype control antibodies. (D) staining for CD14 and IFN-α. (E) Staining with anti-CD14 and an isotype control antibodies. Representing dot plots from one representing patient out of four are shown.

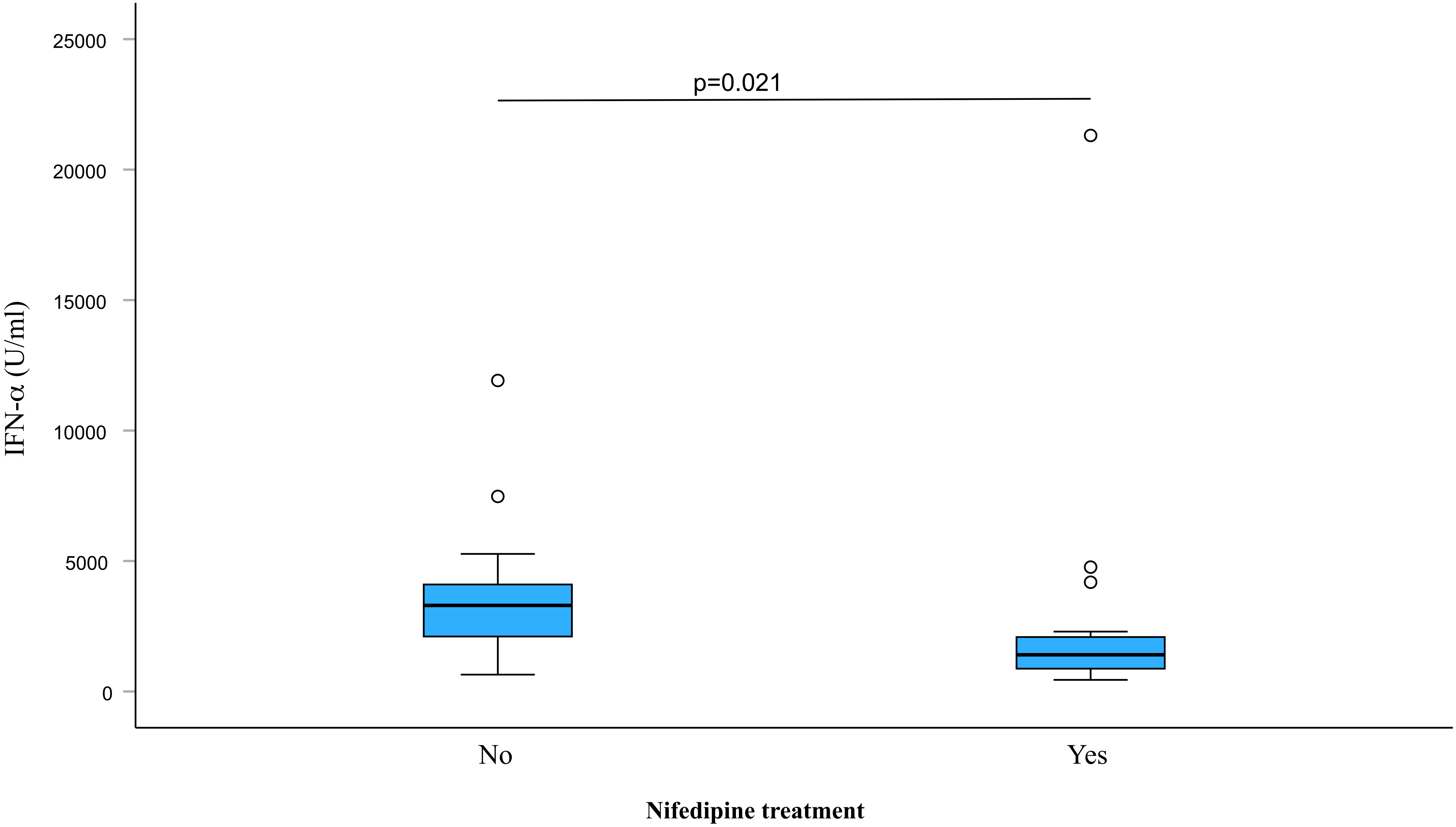
p=0.021
25000
20000
15000
10000
5000
0
IFN-α (U/ml)
No
Yes
Nifedipine treatment

**Supplementary figure 4**

IFN-α production by PBMC isolated from patients with SSc treated or not with the calcium channel blocking drug nifedipine. Peripheral mononuclear cells (PBMC) were stimulated with inactivated herpes simplex virus type I (HSV) for 20h. IFN-α (U/ml) was analyzed in cell culture supernatants by an immunoassay recognizing most IFN-α subtypes. Differences between median values were analyzed with Mann Whitney U test and considered significant if $p \leq 0.05$. The box plots show median (horizontal bars) with interquartile range, minimal and maximal values.

### Supplementary Methods

#### Flow cytometric analysis of surface markers and intracellular IFN-α

The cellular composition of fresh PBMC was analyzed by flow cytometry (FACSCantoII, BD) following surface staining with APC-conjugated anti-BDCA2 (Miltenyi Biotec) and FITC-conjugated anti-CD14 (BD Biosciences) monoclonal antibodies (mAbs).

For intracellular IFN-α detection, PBMC from patients with SSc (n=4) and HC (n=2) were stimulated as described above for 9 hours, with Brefeldin A present during the final three hours. Cells were primed with IFN-α2b to increase intracellular IFN-α production. Surface staining for BDCA2 and CD14 was performed first, followed by fixation and permeabilization with saponin. Intracellular IFN-α was detected by PE-conjugated anti-IFN-α mAb (Miltenyi Biotec) (22). Isotype Ig, fluorochrome-minus-one (FMO), and mock stimulated PBMC served as negative controls.

#### Autoantibody analysis

Information of antibodies to RNP and SSB were collected from patient charts. SSc specific autoantibodies were analysed with a Systemic Sclerosis LIA (Euroimmun, Lübeck, Germany) determining 13 autoantibodies specific for Topoisomerase-1/Scl70, Centromere (CENP-A/B), RNA polymerase III (11kD/155 kD), fibrillarin, NOR90, Th/To, PM-Scl 75/100 kD, Ku, PDGF-R and SSA/Ro52. A semi-quantitative staining intensity grading was obtained by using densitometry (<11 densitometry units negative, 11-25 weakly positive, 26-50 intermediately strong, and >50 strong positive reaction). Analyses and determination of strength of reactions were performed according to the manufacturer's instructions.
